# Strategic validation of variants of uncertain significance in *ECHS1* genetic testing

**DOI:** 10.1101/2022.10.09.22280834

**Authors:** Yoshihito Kishita, Ayumu Sugiura, Takanori Onuki, Tomohiro Ebihara, Tetsuro Matsuhashi, Masaru Shimura, Takuya Fushimi, Noriko Ichino, Yoshie Nagatakidani, Hitomi Nishihata, Kazuhiro R Nitta, Yukiko Yatsuka, Atsuko Imai-Okazaki, Yibo Wu, Hitoshi Osaka, Akira Ohtake, Kei Murayama, Yasushi Okazaki

**Author notes:** These authors contributed equally (Yoshihito Kishita, Ayumu Sugiura).

## Abstract

*ECHS1* is the causative gene for mitochondrial short-chain enoyl-CoA hydratase 1 deficiency. While genetic analysis studies have diagnosed numerous cases with *ECHS1* variants, the increasing number of variants of uncertain significance (VUS) in genetic diagnosis is a major problem. Therefore, we constructed an assay system to verify VUS function. A high-throughput assay using *ECHS1* knockout cells was performed to index these phenotypes by expressing cDNAs containing VUS. The functional validation of VUS identified novel variants causing loss of ECHS1 function. Moreover, we identified cases with functional *ECHS1* defects through multi-omics analysis. We identified a synonymous substitution, p.P163=, and candidate pathogenic variants in the above validation experiments. In summary, this study uncovered new *ECHS1* cases based on VUS validation and omics analysis; these analyses are applicable to functional evaluation of other genes associated with mitochondrial disease.

## Introduction

*ECHS1* encodes short-chain enoyl-CoA hydratase 1 (SCEH), responsible for the degradation of branched chain amino acids and fatty acids (Sharpe & McKenzie, 2018). Abnormalities in valine metabolism particularly impact the pathogenesis of SCEH deficiency (MIM #616277); accumulation of valine metabolites such as S-(2-carbox-ypropyl) cysteine (SCPC) and S-(2-carboxypropyl) cysteamine (SCPCM) derived from methacrylyl-CoA, and S-(2-carboxyethyl) cysteine (SCEC), S-(2-carboxyethyl) cysteamine (SCECM), and 2-methyl-2,3-dihydroxybutyric acid (MDHB), derived from acryloyl-CoA, is often observed (Peters *et al*, 2014). Mutations in *ECHS1* mainly cause Leigh encephalopathy with patients presenting elevated plasma lactate and brain magnetic resonance imaging (MRI) abnormalities (Peters *et al*, 2015; Yamada *et al*, 2015; Haack *et al*, 2015a; Sakai *et al*, 2015; Tetreault *et al*, 2015). In addition, mitochondrial respiratory chain complex abnormalities have also been reported to cause mitochondrial dysfunction in cases with *ECHS1* mutations (Sakai *et al*, 2015; Haack *et al*, 2015a; Tetreault *et al*, 2015). Numerous Japanese cases with *ECHS1* mutations have also been reported (Sakai *et al*, 2015; Haack *et al*, 2015a; Yamada *et al*, 2015; Ogawa *et al*, 2017, 2020). Numerous *ECHS1* variants have been reported, among which, pathogenic variants exist in various genetic regions. A variant frequently reported in Asians is Asn59Ser. The expansion in the clinic of genetic testing has resulted in the rapid accumulation of variants of uncertain significance (VUS). Mitochondrial diseases are no exception, and the VUS number in *ECHS1* is increasing. On the other hand, recent studies have also shown that a valine-restricted diet is effective for cases with *ECHS1* mutations (Yang & Yu, 2020; Sato-Shirai *et al*, 2021). Quick diagnosis is important for early treatment of SCEH deficiency.

Recently, various approaches have been tried to solve VUS. Functional analysis using cultured cells and model organisms is a powerful validation method, providing strong evidence of pathogenicity according to the American College of Medical Genetics and Genomics (ACMG) guidelines (Richards *et al*, 2015). Especially, high-throughput assays and assay methods combining CRISPR/Cas9 and genome-sequencing technologies are being used for VUS verification in cancer-causing genes (Findlay *et al*, 2018; Kweon *et al*, 2020). Although VUS have been validated for *IVD, ACADVL*, and *ACAD9*, all causative genes of inborn errors of metabolism (D’Annibale *et al*, 2021; Xia *et al*, 2021; D’Annibale *et al*, 2022), there is little research on VUS verification in rare diseases.

In this study, we focused on VUS in *ECHS1*, common in many cases of mitochondrial diseases (Kohda *et al*, 2016; Ogawa *et al*, 2017, 2020). In Japan, Tohoku Medical Megabank has a genome project for healthy subjects (Tadaka *et al*, 2019, 2021). In recessive diseases, the causative variant is often present at a certain frequency within a given race. Therefore, in addition to the VUS found in previous our studies of patients with mitochondrial diseases, we conducted a variant validation of rare variants registered in the Japanese Multi-Omics Reference Panel (jMorp).

Here, we constructed an assay system with ATP measurement using cells deficient in the *ECHS1* gene for systematic VUS verification and validation of heterozygotic variants. Furthermore, we found a novel *ECHS1* variant by multi-omics analysis. Coincidentally, the newly found variants were verified by the assay system as being caused by *ECHS1*.

## Results

### Cases with *ECHS1* variants

In previous genomic studies, gene panel sequencing and whole exome sequencing led to the discovery of 15 variants in *ECHS1* cases (Table 1). In Japanese people, the most frequently identified variant was Asn59Ser, followed by Ala2Val (Haack *et al*, 2015b; Sakai *et al*, 2015; Ogawa *et al*, 2020) (Table 1 and Fig. 1). These variants are common in other Asian cases; the allele frequencies of Asn59Ser and Ala2Val are 0.0005769 and 0.0001927 in the gnomAD v3.1.2 East Asian, respectively. Several of these cases also had VUS in ClinVar (as of 20220715, and the same afterhere), such as Met1Val and Leu8Pro (Ogawa *et al*, 2017; Uchino *et al*, 2019). In addition, we also found heterozygous variants in *ECHS1* with conflicting interpretations of pathogenicity (Thr266Ala and Ala278Thr), VUS (Arg272Gln), or unreported (Ala268Thr). All these variants are rare and may be disease-causing, but no experimental verification of the variants has been performed so far. Variants with low allele frequency have been identified, and a number of them have been designated as VUS. Accurate and quick interpretation of these variants is essential for improved diagnosis.

**Table 1.** *ECHS1* variants identified from genomic analysis.

| Patient ID | DNA (NM_004092.4) | Protein(NP_004083.3) |
| --- | --- | --- |
| 0207 | c.5C>T | p.Ala2Val |
| 0207 | c.176A>G | p.Asn59Ser |
| 0255 | c.176A>G | p.Asn59Ser |
| 0255 | c.413C>T | p.Ala138Val |
| 0346ES | c.176A>G | p.Asn59Ser |
| 0346ES | c.476A>G | p.Gln159Arg |
| 0346YS | c.176A>G | p.Asn59Ser |
| 0346YS | c.476A>G | p.Gln159Arg |
| 0376 | c.98T>C | p.Phe33Ser |
| 0376 | c.176A>G | p.Asn59Ser |
| 0536 | c.1A>G | p.Met1Val |
| 0536 | c.5C>T | p.Ala2Val |
| 0775 | c.5C>T | p.Ala2Val |
| 0775 | c.88+2T>C |  |
| 1038ES | c.5C>T | p.Ala2Val |
| 1038ES | c.176A>G | p.Asn59Ser |
| 1038YS | c.5C>T | p.Ala2Val |
| 1038YS | c.176A>G | p.Asn59Ser |
| 1135ES | c.5C>T | p.Ala2Val |
| 1135ES | c.176A>G | p.Asn59Ser |
| 1135YS | c.5C>T | p.Ala2Val |
| 1135YS | c.176A>G | p.Asn59Ser |
| 1553 | c.5C>T | p.Ala2Val |
| 1553 | c.176A>G | p.Asn59Ser |
| 2521 | c.23T>C | p.Leu8Pro |
| 2521 | c.176A>G | p.Asn59Ser |
| 2637 | c.5C>T | p.Ala2Val |
| 2637 | c.176A>G | p.Asn59Ser |
| 2816 | c.23T>C | p.Leu8Pro |
| 2816 | c.176A>G | p.Asn59Ser |

**Figure 1.**
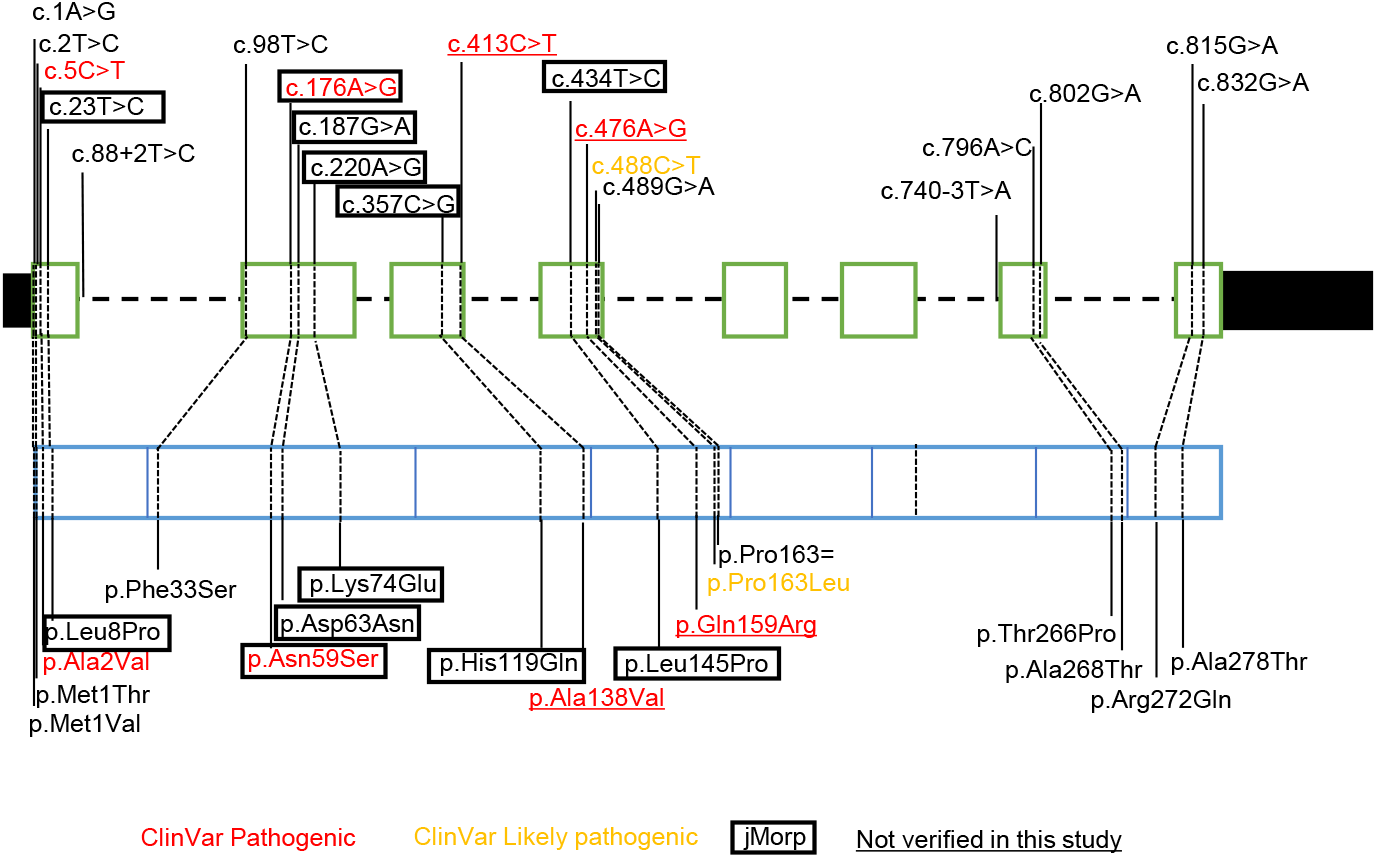
Summary of *ECHS1* variants. Gene structure (top) and corresponding amino acids (bottom) of *ECHS1*. Variants registered r as pathogenic (red) and likely pathogenic (yellow) in ClinVar, and jMorp (square) are shown. The two underlined variants were identified from our genomic analysis study; no experimental VUS verification was performed.

### VUS validation in *ECHS1*

To perform VUS validation of *ECHS1*, we first generated cells deficient in the *ECHS1* gene by targeting *ECHS1* using the CRISPR/Cas9 system in HEK293FT cells. The *ECHS1* KO cell line had an in-frame deletion of 18 bases in exon 2 of the *ECHS1* gene (Fig. 2A). This in-frame deletion resulted in loss of ECHS1 mRNA and protein, as confirmed by qRT-PCR and western blotting (Fig. 2B). We also detected abnormalities in the mitochondrial function in *ECHS1* KO cells. In galactose medium, ATP production is largely dependent on mitochondrial respiration (Robinson *et al*, 1992). Accordingly, we measured ATP levels of wild-type (WT) and *ECHS1* KO cells after incubation in glucose and galactose media. The ratio of ATP levels under galactose and glucose conditions was stable, and no significant difference was observed between *ECHS1* KO and WT cells under these conditions (Fig. 2C). The cellular toxicity of ECHS1 deficiency is due to accumulation of intermediates in valine metabolism. That explains why a valine-restricted diet is recommended to patients harboring *ECHS1* pathogenic variants (Sato-Shirai *et al*, 2021). Consistently, valine addition to the galactose medium lead to further reduced ATP level in *ECHS1* KO cells, in a dose-dependent manner (Fig. 2D). These results suggest that the abnormal mitochondrial function in *ECHS1* KO cells results from the abnormal valine metabolism, as shown in human patients, making it a suitable model to validate VUS of *ECHS1*. Accordingly, we performed a functional verification of VUS using *ECHS1* KO cells expressing VUS.

**Figure 2.**
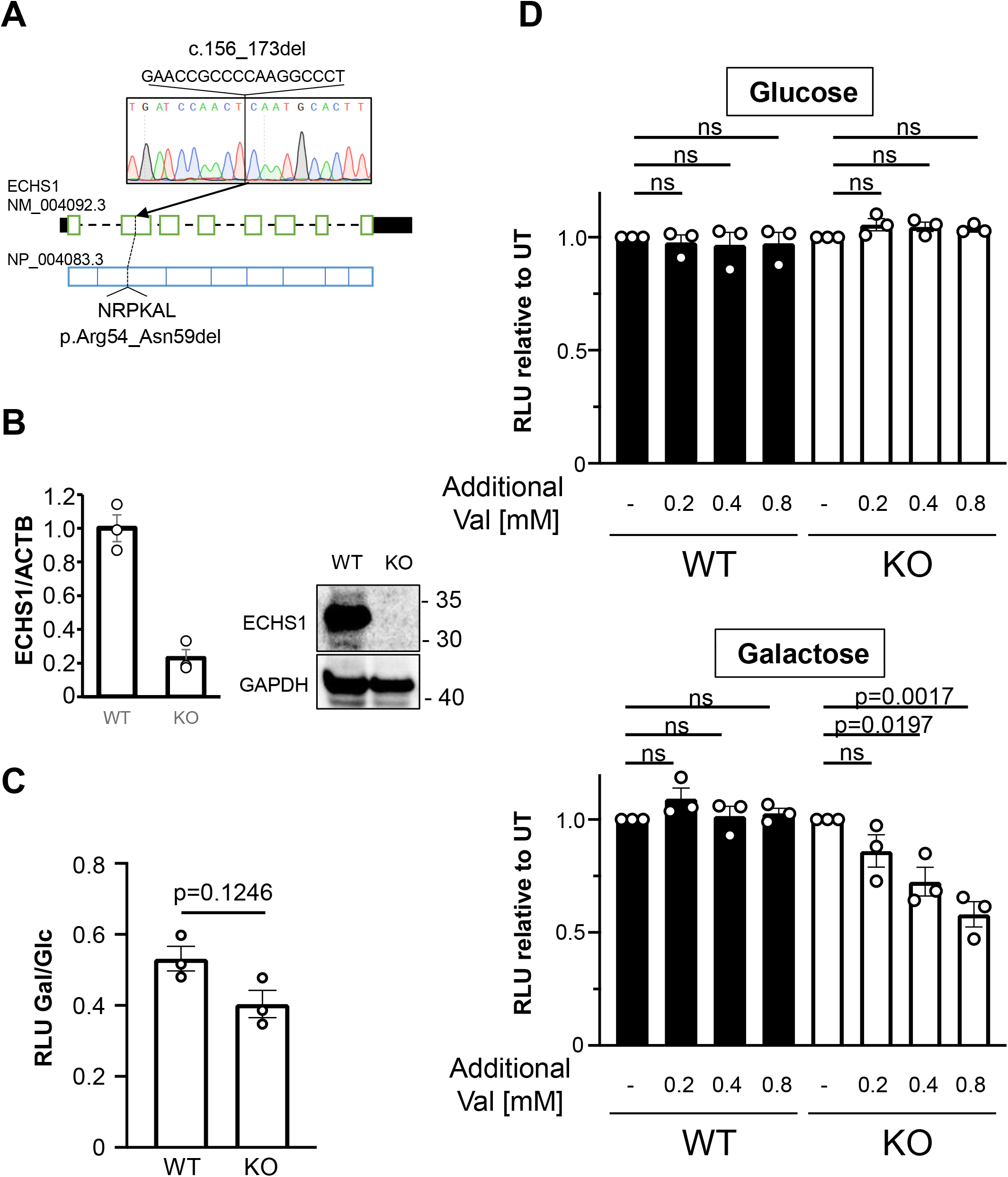
Characterization of *ECHS1* KO cells. A. Genomic analysis of *ECHS1* KO cells by Sanger sequencing. B. *EHCS1* expression levels in WT and *ECHS1* KO HEK293FT cells quantified by qRT-PCR. Whole cell extracts from WT and *ECHS1* KO HEK293FT cells analyzed by immunoblotting using the indicated antibodies. Single and double asterisks indicate uncleaved and cleaved forms, respectively. C. ATP assay of WT and *ECHS1* KO HEK293FT cells cultured in glucose or galactose medium. The graph shows the ATP level in galactose medium divided by galactose. D. ATP assay of WT and *ECHS1* KO HEK293FT cells treated with the indicated L-valine concentrations. Bar graphs represent the average ATP level in each condition from three biological independent experiments. Error bars, ±SEM. Statistical analysis was performed using ANOVA followed by Dunnet’s test. RLU, relative luciferase unit.

First, variants identified from previous genomic studies and from jMorp were selected for functional validation. The targets were 15 variants identified by our genome analysis and six rare variants registered in jMorp (4.7KJPN released on 20190902; Fig. 1 and Table S1). The Leu8Pro and Asn59Ser variants were found in our patients as well as registered in jMorp (4.7KJPN). Other four variants were not identified in our genome analysis of mitochondrial disease patients. ECHS1 is a mitochondria-localized protein with a mitochondrial targeting sequence (MTS) at the N-terminal. Four variants were located at the translation initiation site and the MTS. We compared *ECHS1* KO cells transfected with an empty vector and *ECHS1* WT cDNA to *ECHS1* KO cells transfected with an *ECHS1* gene including VUS.

Exogenously expressed ECHS1 shows uncleaved and cleaved forms (Fig. 3A, single and double asterisks, respectively). Some variants were dominantly expressed in an uncleaved form. Further, we found a significant decrease in the expression level of variants in the MTS and at leucine 145 (Fig. 3A).

**Figure 3.**
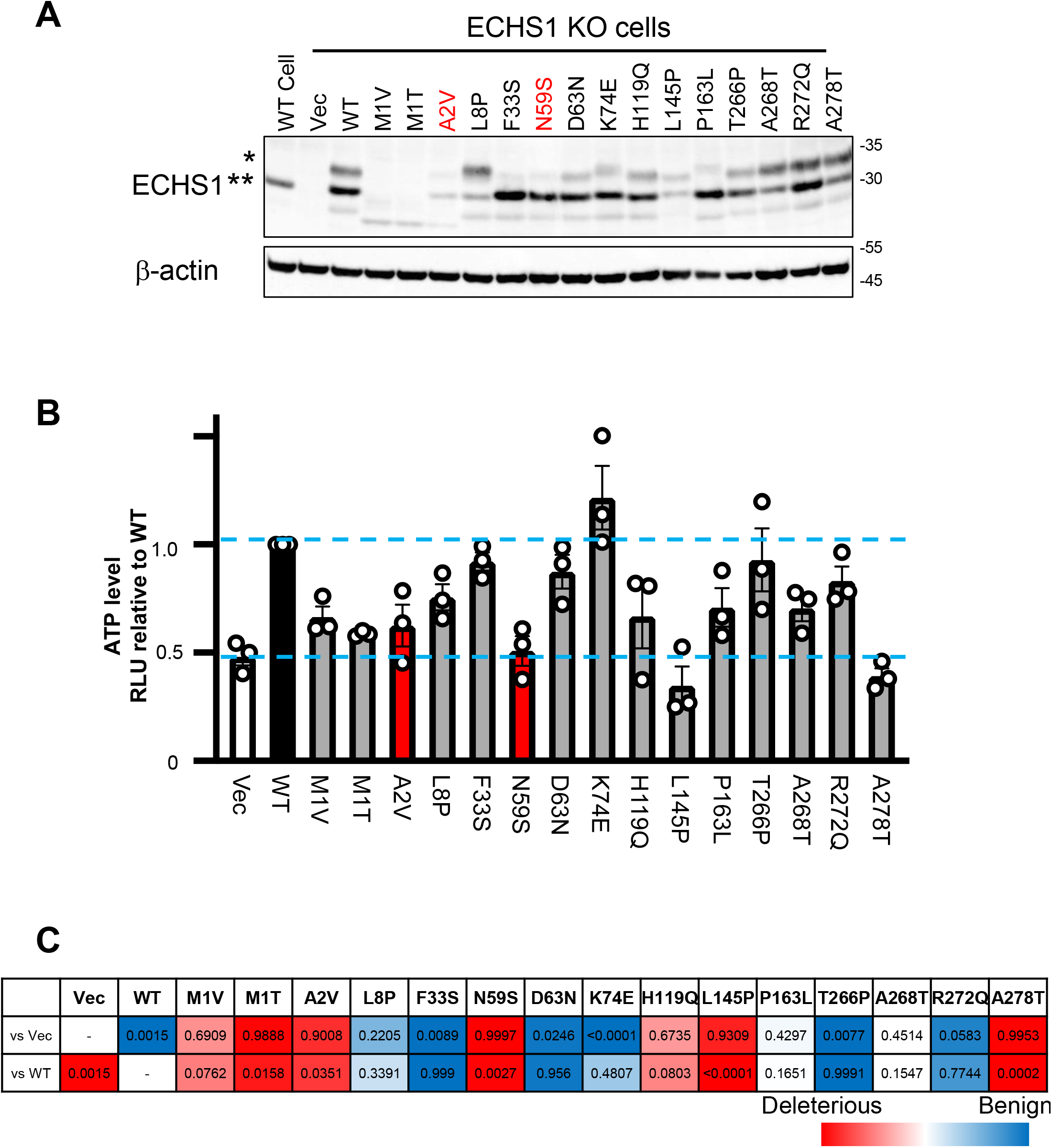
Validation of *ECHS1* VUS. WT and *ECHS1* KO HEK293FT cells transfected with the indicated expression vectors subjected to immunoblotting analysis (A) and ATP assay (B). A. Whole cell extracts were analyzed by immunoblotting using the indicated antibodies. Single and double asterisks indicate uncleaved and cleaved forms, respectively. B. ATP assay 4 days after treatment with L-valine (0.8 mM). Bar graphs represent the average ATP level in each condition from three biological independent experiments. Error bars, ±SEM. Red bars, pathogenic variants. Gray bars, VUS. RLU, relative luciferase unit. C. Statistical analysis of Figure B using ANOVA followed by Dunnet’s test. The color scale shows the P value compared with vector (vs Vec, statistically different red to blue) and WT *ECHS1* (vs WT, statistically different blue to red).

Next, we examined VUS functions in the mitochondria. *ECHS1* WT expression restored ATP levels in *ECHS1* KO cells (Fig. 3B). The ATP expression of KO cells transfected with each variant was compared with that for KO cells transfected with empty vector or transfected with WT vector; changes in ATP levels were evaluated by one-way ANOVA followed by Dunnet’s test (Fig. 3C). In addition to Ala2Val and Asn59S, reported as pathogenic variants, the two VUS with altered start codons failed to restore ATP levels probably due to significantly reduced protein expression (Fig. 3B). A marked down-regulation in protein levels was observed, with Leu8Pro showing a significant reduction in the lower band considered to correspond to mature ECHS1 (Fig. 3A). His119Gln, Pro163Leu, Ala268Thr, and Ala278Thr variants had higher expression levels, but failed to fully improve ATP levels equivalent to WT, suggesting that these variants were deleterious (Fig. 3B).

Although Leu8Pro and Phe33Ser were reported as pathogenic variants (Kohda *et al*, 2016; Uchino *et al*, 2019), ATP assays demonstrate a mild functional decline. Since Leu8Pro and Phe33Ser were identified as a compound heterozygote with Asn59Ser, a nonfunctional variant (Fig. 3B), we hypothesized that Leu8Pro and Phe33Ser might be mild deleterious variants capable of restoring KO cells but not KO cells expressing highly toxic variants. To test this hypothesis, we reproduced compound heterozygous genotypes by transfecting *ECHS1* KO cells with two different variants (Fig. 4A). The ATP assay demonstrated that WT restored KO cells upon co-transfection with Asn59Ser, whereas, as expected, all three variants tested did not show recovery (Fig. 4B and C). Our system is valuable to validate the pathogenic nature of the compound heterozygous state.

**Figure 4.**
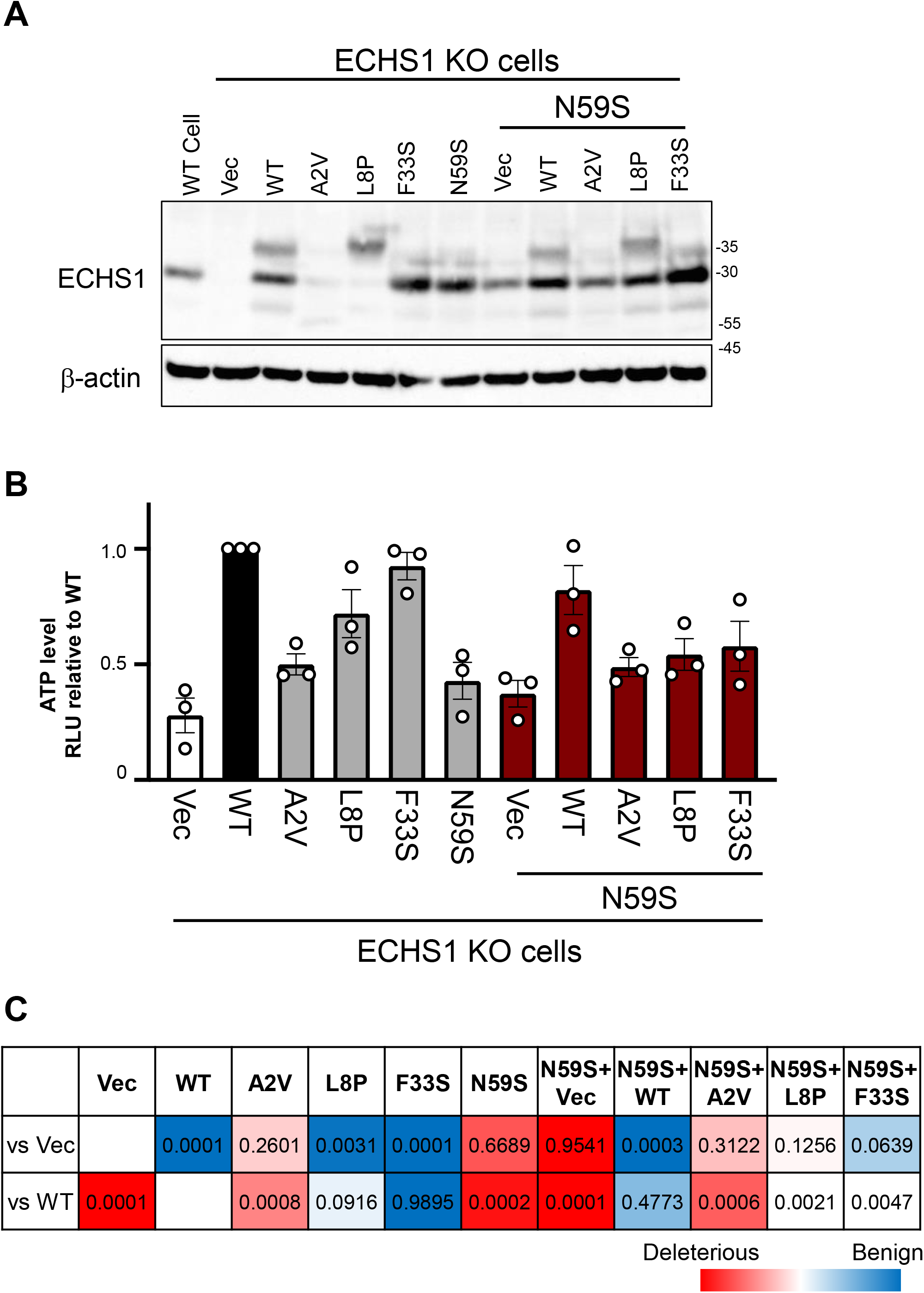
Validation of ECHS1 VUS compound hetero. A. Whole cell extracts from WT and *ECHS1* KO HEK293FT cells transfected with the indicated expression vectors analyzed by immunoblotting using the indicated antibodies. B. ATP assay of *ECHS1* KO HEK293FT cells expressing the indicated expression vectors treated with additional L-valine (0.8 mM) for four days. Bar graphs represent the average ATP level in each condition from three biological independent experiments. Error bars, SEM. RLU, relative luciferase unit. C. Statistical analysis of Figure B using ANOVA followed by Dunnet’s test. The color scale shows the P value compared with vector (vs Vec, statistically different red to blue) and WT *ECHS1* (vs WT, statistically different blue to red).

### Validation of ECHS1 variants from omics analysis

In case 1, a Thr266Pro variant was identified; in case 2, an Ala268Thr variant was identified; in cases 3 and 4, a Ala278Thr variant was identified; in case 5, an Ala278Val variant was identified (Table 2, Fig. S2). In addition, these cases also had a Pro163= variant, identified in a Samoan family and a very frequent variant (0.01156) in gnomAD v3.1.2 East Asian populations, and suggested to exhibit splicing abnormalities (Simon *et al*, 2021). In the present study, RNA-seq analysis was performed on case 1 and case 4. Then, we identified sequence reads showing exon skipping as consequence of the Pro163= variant (Fig. 5A). To examine whether exon skipping actually increased in cases with Pro163=, we plotted RNA-seq counts of the *ECHS1* gene and the number of detected reads showing exon skipping (Fig. 5B). We analyzed 26 RNA-seq data, including case 1 and case 4, as well as one case with heterozygous Pro163=; exon skipping increased in these samples. In addition, after counting the allele numbers at the Thr266Pro and Ala278Thr variant positions showed a bias in allele expression (Fig. 5C). This indicates that the significantly reduced expression of alleles with Pro163= in case 1 and case 4 fibroblasts. Furthermore, proteome analysis in case 1 fibroblasts confirmed a significant decrease in ECHS1 expression (Fig. 6A). In addition, protein expression in case 1 fibroblasts was confirmed by sodium dodecyl sulfate polyacrylamide gel electrophoresis (SDS-PAGE) followed by Western blot (WB), revealing a marked decrease in protein expression (Fig. 6B). Furthermore, previous studies experimentally demonstrated that case 4 leads to decreased ECHS1 expression and enzymatic activity, as well as accumulation of intermediate products of valine metabolism (Kuwajima *et al*, 2021). We here concluded that and non-synonymous substitution combinations are disease-causing in these cases.

**Table 2.**
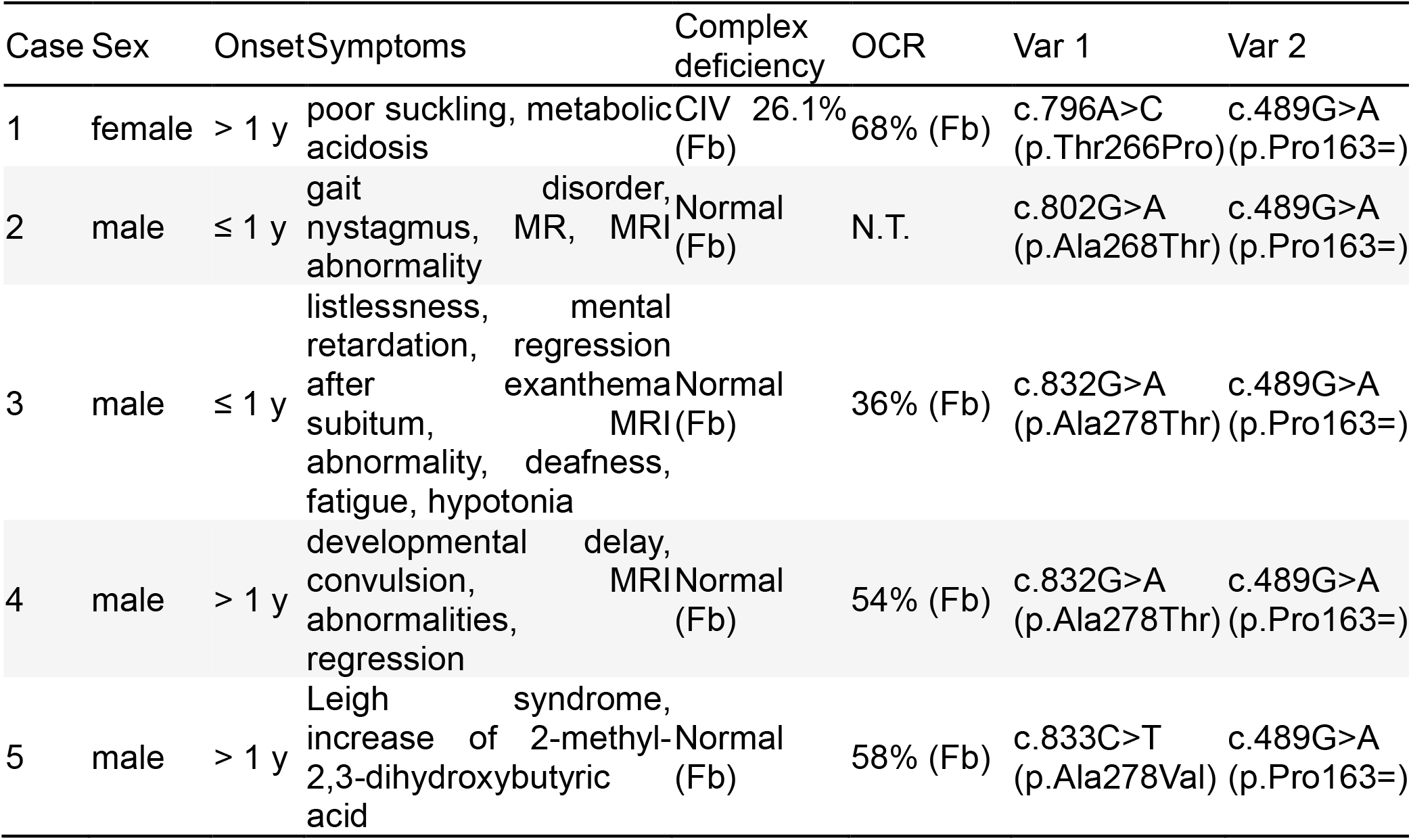
Patient summary of cases with *ECHS1* variants. Fb: Fibroblast, CIV: Mitochondrial respiratory chain complex IV, OCR: oxygen consumption rates. Complex enzyme activity was defined by <40% decrease. For OCR, a value <71.6% was used as diagnostic criterion.

**Figure 5.**
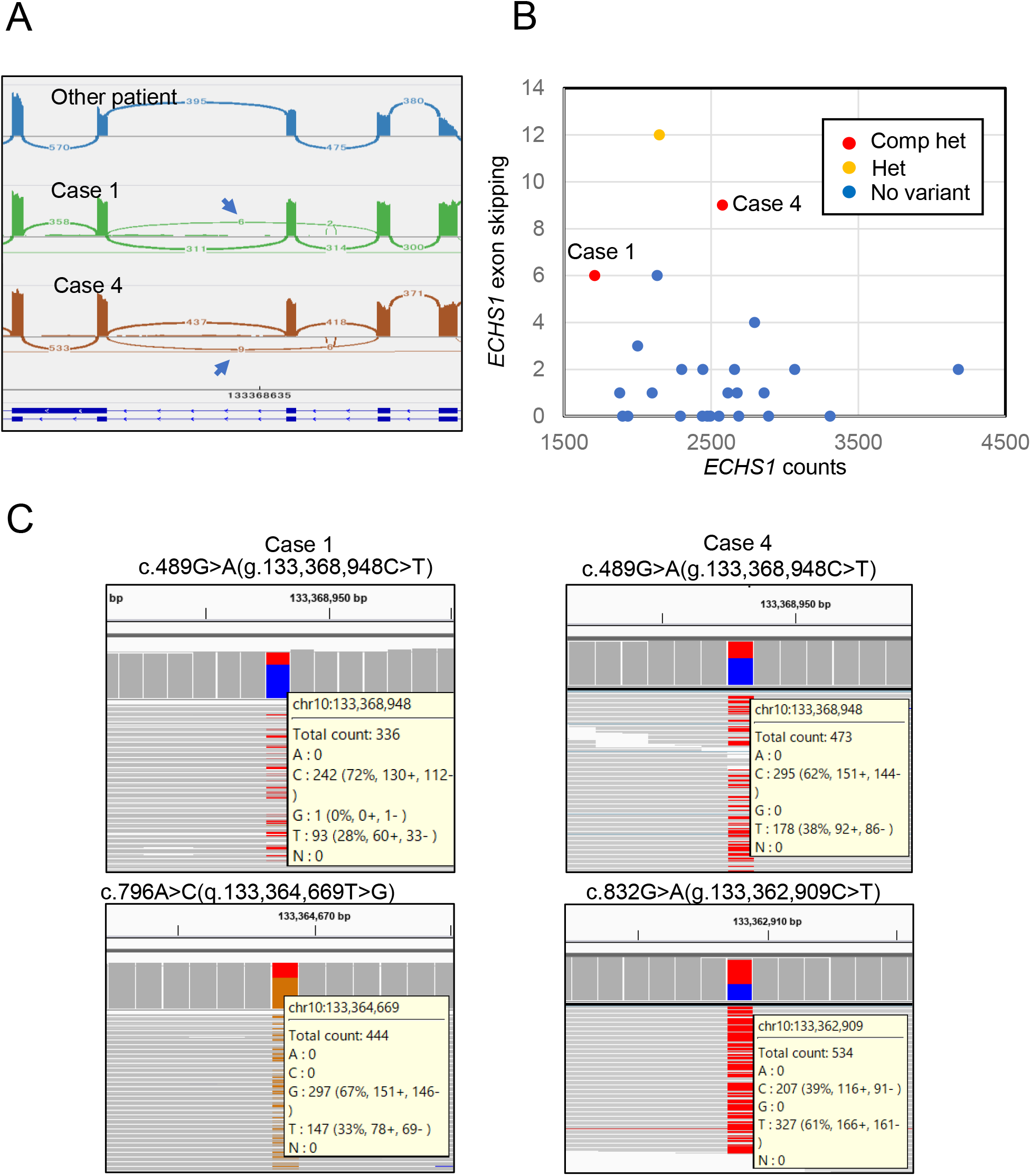
Abnormal splicing and allele-biased gene expression. A. RNA-seq data from two patients with c.489G>A (p.P163=) showing reads suggestive of splicing abnormalities in exons with c.489G>A. B. Read counts of the *ECHS1* gene are plotted on the horizontal axis and the number of detected exon skipping is plotted on the vertical axis. Gene counts were calculated by STAR, and the number of exon skipping was extracted from the Sashimi plot data. C. Ratio of c.489G>A and c.796A>C and c.832G>A variants on the IGV viewer. In two cases, the allele expression with c.489G>A was decreased.

**Figure 6.**
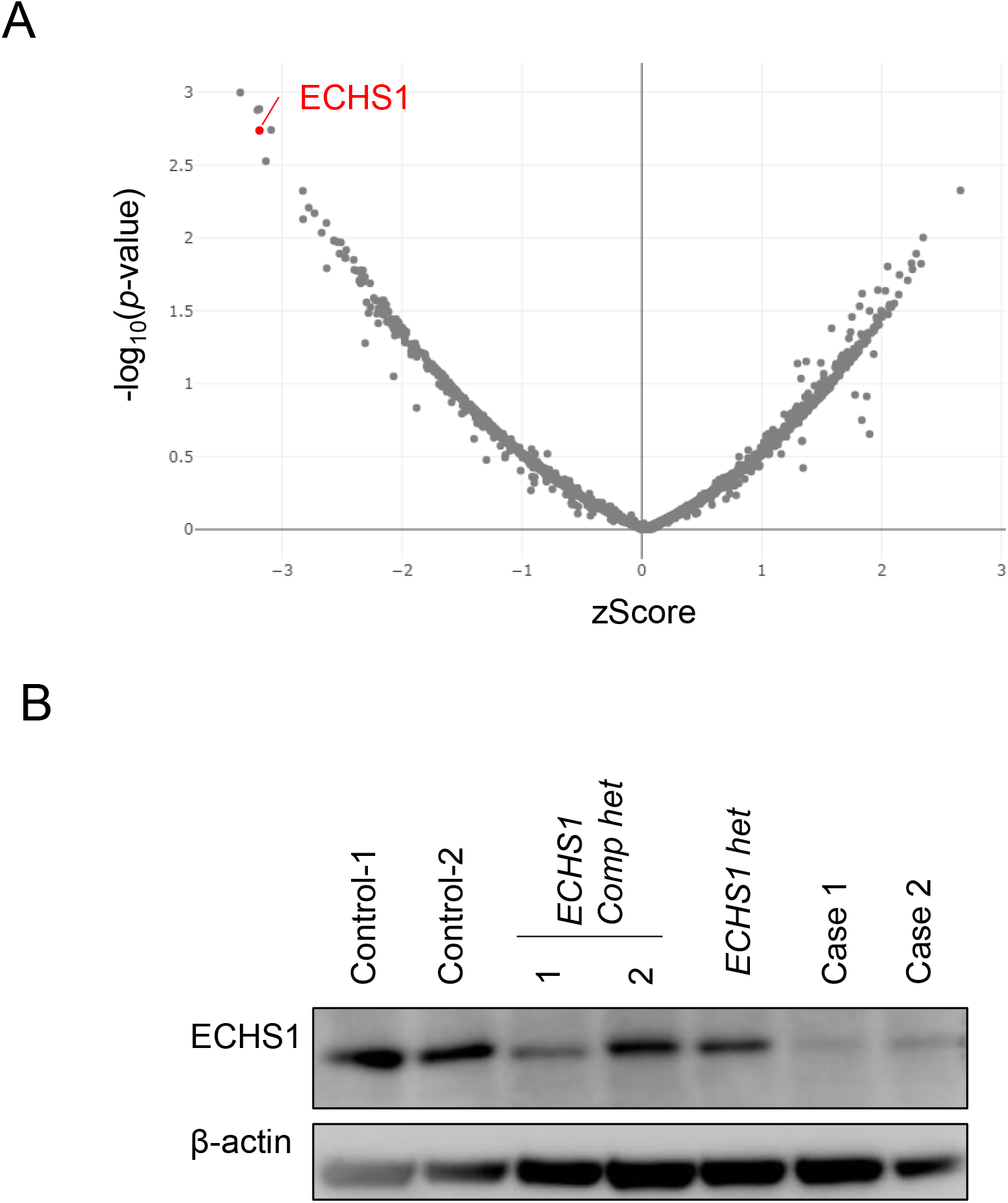
ECHS1 protein status in the patient with Pro163= and Ala278Thr. A. OUTRIDER analysis illustrating protein expression in a volcano plot. ECHS1 was detected as a protein with a large decrease in expression. B. Western blotting for ECHS1 in patients with *ECHS1* mutations and controls. Compared to previously reported ECHS1 cases, case 1 showed significantly reduced ECHS1 expression. β-actin was detected as a loading control.

## Discussion

*ECHS1* is one of the most frequent genes found in patients with mitochondrial diseases. In addition to our genetic study(Haack *et al*, 2015b; Ogawa *et al*, 2017, 2020), *ECHS1* has been reported as a frequent cause of mitochondrial disease in genetic studies of Leigh encephalopathy in Asian countries such as China(Stenton *et al*, 2022) and Korea(Lee *et al*, 2020). The length of the coding region is 873 bp. About 250 variants have been reported in ClinVar, of which about 40 are pathogenic/likely pathogenic and nearly 80 have been registered as VUS or with conflicting interpretations of pathogenicity. However, the molecular mechanism by which *ECHS1* variants cause disease is poorly understood. In this study, we established a system to functionally validate *ECHS1* variants, finding some potential pathogenic variants.

Resolving VUS is a major challenge in various diseases, because VUS continue to accumulate under the circumstances where genome analysis is becoming more common. Genome sequence projects in healthy individuals have revealed allele frequencies in various races. The accumulation of genome analysis information is expected to lead to the discovery of novel pathogenic variants, since rare variants also found in healthy individuals can be pathogenic. Against this background, it may be possible to make a proactive evaluation of variants not yet been associated with disease. Based on the above, we could identify novel pathogenic variants in this study. Furthermore, the combination of RNA-seq, proteomics and conventional genomic analysis enabled a reliable diagnosis.

*ECHS1* is a nuclear-encoded gene transported to mitochondria by the MTS(Burgin & McKenzie, 2020). MTS locates at the N-terminus and is cleaved in mitochondria by mitochondrial proteases(Vaca Jacome *et al*, 2015). There are three points involved in pathogenicity: 1) expression, 2) location, and 3) function. It was reported that ECHS1 is degraded through ubiquitin-proteasome pathway in cancer cells (PMID: 34615856). As shown in Fig. 2A, Met1Ala, Met1Thr, and Ala2Val were weakly expressed. Consistently, these variants failed to restore ATP decrease in *ECHS1* KO cells. Since inhibition of proteasome degradation by MG132 had no effect on the protein level of all *ECHS1* variants tested in this study (Fig. S1), these substitutions may alter the expression level at the transcriptional or translational level. The Leu8Pro variant was detected dominantly in the uncleaved form (Fig. 3A), suggesting mis-targeting to mitochondria. Leu145Pro was also weakly expressed, but increased expression with higher amount of transfection did not restore. As with Asn54Ser, Leu145Pro is a functionally disrupted variant. Interestingly, Leu145Pro, a variant reported only in jMorp and not previously identified in patients with the disease, had a high molecular weight. It is expected to be reported in Japanese case with this variant. We propose that those who identify variants in ECHS1 by genetic test will refer to our variant evaluation study as evidence and that this will lead to a solution to the cause of the disease in the future.

Leu8Pro and Phe33Ser, considered “Likely benign” variants from VUS validation experiments (Fig. 3B and C), were validated by co-expressing with the Asn59Ser variant (Fig. 4B and C). By validating the assumption of complex heterozygosity in combination with a pathogenic variant, we could obtain data accurately reflecting the functional evaluation of hypomorphic variants. The assay was effective for variants in the borders, for which very subtle validation results were obtained. Despite its higher experimental complexity, the assay can be performed while maintaining the conventional throughput, and the validation targets can be expanded. In our reported *ECHS1* mutant cases, the most common combination of is Asn59Ser and Ala2Val. However, since Asn59Ser has a higher allele frequency than Ala2Val, cases homozygous for Asn59Ser should be more frequent; however, no patients homozygous for Asn59Ser have been found. This suggests that Asn59Ser may be so harmful that in homozygosity it may result in severe developmental abnormalities in the prenatal period. The high number of Asn59Ser and Ala2Val combinations may be due to Ala2Val having a smaller functional loss than Asn59Ser, as shown experimentally in this study. Asn59Ser is only viable in a compound heterozygous state with less toxic variants. The combination with milder variants such as Leu8Pro and Phe33Ser, verified in this study, might allow normal development until birth. In other words, this combined assay could first accurately indicate functional abnormalities for variants with abnormalities intermediate between normal and completely defective, being a very effective and essential method for variant evaluation. Mostly, in silico predictions and experimental validation of most variants are comparable. On the other hand, silico predictions for some variants differ from our validation experiments; therefore, validation experiments are important for such variants. For example, Ala2Val, already been reported as pathogenic, was not highly damaging in in silico predictions (Table S1). However, the validation results suggest that although the experimental data indicate an effect on protein gene expression itself, the functional effect may not be as significant. A similar trend was observed for Leu8Pro, expected to have a significant effect on protein localization but relatively little effect on protein function. His119Gln was also rated mostly tolerant or benign in silico, but experimental validation suggested that the variant affects gene function. The major achievement of this study is providing quantitative experimental validation data for each variant on the same functional platform. We are expanding the validation of other genes; in particular, VUS validation of genes involved in respiratory chain complex I confirmed this to be a reproducible and efficient analysis.

In this validation, we focused on variants specific to Japanese and Asian populations. Naturally, this should be further expanded to include variants from other ethnicities. Our assay system is based on a very simple method and can easily be expanded to other VUS, by including gnomAD and ClinVar variants. In the present study, VUS was verified for jMorp (4.7KJPN) variants, but since then, the jMorp data has been updated and the number of registrations has increased. While planning our validation experiment, the variant data of 4.7K JPN were registered, from which we extracted variants with very low allele frequencies (Table S2). However, there are now 38K JPN, with 20 more registrations from our validated variants. The data is being updated at an accelerated pace, with 14KJPN in 2021 and 38KJPN in 2022. Given the constant updates to the database, it is important to verify these variants in the future.

ECHS1 protein loss is considered to have a threshold of 30–40% for disease(Simon *et al*, 2021). This is shown by the analysis with Pro163= and Ala278Thr from the study of Simon et al. Those homozygous for Pro163= did not develop the disease, despite showing a protein expression around 40%. On the other hand, the combination of Pro163= and Ala278Thr showed <30% protein expression and developed the disease. Ala278Thr is considered highly toxic, as it was found to be deleterious in the VUS validation experiment. Although Ala278Val has not been validated, we think it likely to be as deleterious as Ala278Thr since many scores in silico also indicated damaging effects. For Thr266Pro, no significant decrease in ATP levels was observed in VUS validation. However, abnormalities at the protein level were evident from the proteome and immunoblotting. VUS expression experiments with the Thr266Pro variant also showed reduced ECHS1 expression (Fig. 3A). Considering these results, Thr266Pro might have less functional loss and more impact on protein expression. In addition, the forced expression system showed a protein decrease not expected to significantly reduce ATP levels. In protein expression validation (Fig. S1), the amount of mature ECHS1 synthesized from Thr266Pro was lower than that of immature ECHS1, suggesting that this variant has a significant effect on protein expression and maturation. For Ala268Thr, a mild decrease in ATP levels was observed. Since the value of Ala268Thr was similar to that of Pro163Leu, reported as likely pathogenic in ClinVar, Ala268Thr was considered to have an effect on protein function. Case 2 with the Ala268Thr variant had later onset and milder symptoms than other ECHS1 cases, suggesting that the variant itself has a milder effect. Pro163=, despite its high allele frequency [0.01156 in gnomAD v3.1.2 East Asian, 0.00789 in the jMorp (38KJPN)], is a possible causative variant; its combination with other pathogenic variants leads to disease development. We found four cases with Pro163= and other rare variant in this study. There have been very few reports of such a high frequency variant in studies of mitochondrial diseases; this variant might be responsible for the increased frequency of cases with ECHS1 mutations. However, variants without amino acid substitutions have been overlooked as benign in previous genetic diagnosis. Given the high potential number of patients with Pro163=, it is expected that the number of cases with Pro163= will further increase because of our findings.

Our VUS verification system has some limitations. Even pathogenic variants may be missed in this experimental system. In fact, Pro163Leu, registered as likely pathogenic in ClinVar, showed a mild score. Thus, variants with intermediate ratings could be pathogenic. In such cases, it would be necessary to validate them using an assay system that assumes compound heterozygosity. Moreover, since this is a forced expression system, variants that cause protein stability or splicing abnormalities may be missed. To extract these variants, other assay systems and functional experiments using patient specimens are required.

## Materials and methods

### Cell culture and knockout cell generation

Cells were cultured at 37°C and 5% CO_2_ in Dulbecco’s modified Eagle’s medium (DMEM with 4.5 g/L glucose; Nacalai Tesque) supplemented with 10% fetal bovine serum and 1% penicillin–streptomycin.

Single guide RNAs (sgRNAs) were designed using CRISPRdirect software (Naito *et al*, 2015).he target sequence was as follows: 5’-GGGCCTTGGGGCGGTTCAGT-3’. gRNA oligonucleotides were inserted into a pSpCas9(BB)-2A-Puro (PX459) V2.0 (Addgene 62988) plasmid as previously described(Ran *et al*, 2013). HEK293FT cells were transfected with PX459 including *ECHS1* targeted sgRNA. Cells were selected using 2 µg/mL puromycin and single cells were isolated. Genomic DNA was extracted from isolated cells, and sgRNA target sites were amplified using KOD FX Neo (Toyobo). Primer sequences are as follows: 5’-CCCATGACCGTCTTCACTCG-3’ and 5’-ACATCCCTTCCCCCACTCTC-3’. PCR products were purified and directly sequenced.

### Vector construction

cDNA of *ECHS1* (NM_00492) WT and VUS were synthetized and inserted into pCDNA3.1 Lifect-EGFP (Addgene 67303) with HindIII and XbaI sites by GENEWIZ/Azenta.

### ATP assay

HEK293FT WT or *ECHS1* knockout cells were seeded in a collagen-coated 96-well plate (354650, Corning, AZ, USA) at 1 × 10^4^ cells/well with growth medium containing 25 mM glucose. For VUS validation, *ECHS1* knockout cells were transfected with 20 ng of expression vectors encoding WT or *ECHS1* variants. One day after plating or transfection, the medium was replaced with 25 mM glucose or 10 mM galactose medium supplemented with dialyzed 10% fetal bovine serum (04-011-1A, Biological industries, KibbutzBeit-Haemek, Israel) and L-valine (13046-62, Nacalai Tesque, Kyoto, Japan). Four days after culture in galactose or glucose medium, the ATP content was measured using the CellTiter-Glo® Luminescent Cell Viability Assay kit (Promega) with a VICTOR Nivo multimode microplate reader (PerkinElmer, MA, USA).

### RNA-seq

RNA was purified from fibroblasts by the Maxwell RSC simplyRNA Cells Kit and a Maxwell RSC Instrument (Promega). After quality check by Agilent 2100 and Qubit 2.0, the mRNA was enriched using oligo(dT) beads and rRNA removed using the Ribo-Zero kit. The mRNA was fragmented randomly by adding fragmentation buffer; then, cDNA was synthesized by using the mRNA template and random hexamers primers, followed by addition of a custom second-strand synthesis buffer (Illumina), dNTPs, RNase H and DNA polymerase I to initiate second-strand synthesis. Second, after a terminal repair, A ligation, and sequencing adaptor ligation, the double-stranded cDNA library was completed through size selection and PCR enrichment. Sequencing was performed using 150-bp paired-end reads on a NovaSeq6000 (Illumina). Fastq files were aligned to the GRCh38/hg38 genome by STAR. Gene read counts were quantified by STAR quantMode GeneCounts function. The aligned BAM files were loaded into the Integrated Genomics Viewer and visualized using a Sashimi plot for mRNA splicing analysis.

### qRT-PCR

RNA was isolated from culture fibroblasts using FastGene RNA Basic Kit. The isolated RNA was reverse-transcribed to cDNA using ReverTra Ace® qPCR RT Master Mix (TOYOBO) according to manufacturer’s instructions. The synthesized cDNA was used as a template for qRT-PCR in a 7500 Fast Real-Time PCR System (Thermo Fisher Scientific) using THUNDERBIRD® Probe One-step qRT-PCR Kit (TOYOBO). Primer sequences are as follows: ECHS1-F, 5’-GTCTTCAGGGCCTGGTTGAG-3’, ECHS1-R, 5’-CTGTGCAAACTGGGCCTTCT-3’, ACTB-F, 5’-GCGAGAAGATGACCCAGATC-3’, ACTB-R, 5’-GGATAGCACAGCCTGGATAG-3’.

### Sanger sequencing

The *ECHS1* variants of patients and family members were sequenced by Sanger sequencing. PCR products were directly sequenced using BigDye v3.1 Terminators (Applied Biosystem, ThemoFisher Scientific) or SupreDye v3.1 reagent (Edge BioSystems) and ABI 3130XL (Applied Biosystems, ThemoFisher Scientific).

### Proteome

Samples from fibroblasts were prepared as described previously (Borna *et al*, 2019). The samples were measured in both data-dependent and data-independent modes performed on the Q-Exactive Plus mass spectrometer (Thermo Fisher Scientific) as previously described (Borna *et al*, 2019). Finally, 5,979 proteins were detected in 16 samples including two healthy controls and 14 mitochondrial disease patients. Then, outlier protein expression analysis was performed using OUTRIDER (Brechtmann *et al*, 2018).

### Immunoblotting analysis

SDS–PAGE and western blot were performed as previously described (Kohda *et al*, 2016). To isolate mitochondria, cell pellets were suspended in mitochondria isolation buffer A (220 mM mannitol, 20 mM HEPES, 70 mM sucrose, 1 mM EDTA, pH 7.4, 2 mg/mL bovine serum albumin, 1× protease inhibitor cocktail) and homogenized with 20 strokes on ice. Homogenates were separated into cytosolic and nuclear fractions after centrifugation at 700 g for 5 min at 4°C. The supernatants were centrifuged at 10,000 g for 10 min at 4°C. Mitochondrial pellets were rinsed twice with mitochondria isolation buffer B (220 mM mannitol, 20 mM HEPES, 70 mM sucrose, 1 mM EDTA, pH 7.4, 1× protease inhibitor cocktail). Then, mitochondrial protein levels were determined using a bicinchoninic acid assay. For SDS–PAGE analyses, enriched mitochondria were solubilized in RIPA buffer (Nacalai Tesque, Japan) and denatured for 5 min at 95°C. Prepared samples were separated by electrophoresis on 10% or 15% SDS–PAGE gels, depending on the size of the detected protein. Each antibody was obtained as follows; ECHS1 (11305-1-AP, Proteintech, IL, USA), GAPDH (G9545, Sigma-Aldrich), beta-actin (A5441, Sigma-Aldrich).

### Ethics statement

The studies were approved by the regional Ethics Committees at Juntendo University, Saitama Medical University, and Chiba Children’s Hospital, Kindai University. We obtained written informed consent from the parents. All methods were performed in accordance with relevant guidelines and regulations.

### Statistics

Data are expressed as the mean ± SEM. The statistical significance of differences was determined by one-way ANOVA followed by Dunnet’s test using Prism 9 (GraphPad Software Inc., CA, USA).

## Data availability

Raw data are available from the corresponding author upon reasonable request. ECHS1 knockout cells can also be distributed. Some genomic information that could be used to identify individuals cannot be shared due to ethical restrictions.

## Acknowledgments

We thank the family for their participation in the research presented here and the Laboratory of Molecular and Biochemical Research, Biomedical Research Core Facilities, Juntendo University Graduate School of Medicine, and Kasumi Kanai for technical assistance. This work was supported by a grant for the Practical Research Project for Rare/Intractable Diseases from AMED to H.O., K.M., Y.O. and A.O. (Fund ID: JP21im0210625, JP21ek0109511, JP22ek0109485, JP22ek0109468, JP22gk0110038, JP19ek0109273), Program for Promoting Platform of Genomics based Drug Discovery to Y.O. (Fund ID: JP22kk0305015), the Research Center Network for Realization of Regenerative Medicine (The Acceleration Program for Intractable Diseases Research utilizing Disease-specific iPS cells, JP21bm0804018), and JSPS KAKENHI JP19H03624 to Y.O. and JP20H03648 to H.O.

## Author contributions

YK, AS and YO wrote the manuscript. YK, AS, TK, TE, TM, MS, NI, YN, HN, YY and YW performed the experiments. YK, AS, TK, TE, TM, MS, NI, YN, HN, YY, AIO, YW and YO analyzed the data. TK, TE, TM, MS, TF, HO, AO and KM acquired clinical information. YK, AS, TF, KRN and AIO did bioinformatics and statistical analysis. YK, AS and AO supervised the study. All authors discussed the results and commented on the manuscript.

