## Supplementary information (Figures S1&S2, Tables S1&S2) for "Strategic validation of variants of uncertain significance in *ECHS1* genetic testing"

**Title**

**Running title**

VUS validation in *ECHS1* genetic testing

**Authors**

Yoshihito Kishita[1,2,12], Ayumu Sugiura[2,12], Takanori Onuki[3], Tomohiro Ebihara[4], Tetsuro Matsuhashi[3], Masaru Shimura[3], Takuya Fushimi[3], Noriko Ichino[2], Yoshie Nagatakidani[1], Hitomi Nishihata[1], Kazuhiro R Nitta[2], Yukiko Yatsuka[2], Atsuko Imai-Okazaki[2], Yibo Wu[5,6], Hitoshi Osaka[7], Akira Ohtake[8,9], Kei Murayama[3,10], Yasushi Okazaki[2,11,*]

**Affiliations**

[1]Department of Life Science, Faculty of Science and Engineering, Kindai University, Osaka, Japan

[2]Diagnostics and Therapeutics of Intractable Diseases, Intractable Disease Research Center, Juntendo University, Graduate School of Medicine, Japan

[3]Department of Metabolism, Chiba Children's Hospital, Chiba, Japan

[4]Department of Neonatology, Chiba Children's Hospital, Chiba, Japan

[5]University of Geneva, Geneva, Switzerland.

[6]YCI Laboratory for Next-Generation Proteomics, RIKEN Center of Integrative Medical Sciences, Kanagawa, Japan

[7]Department of Pediatrics, Jichi Medical University, Tochigi, Japan

[8]Department of Pediatrics & Clinical Genomics, Faculty of Medicine, Saitama Medical University, Saitama, Japan

[9]Center for Intractable Diseases, Saitama Medical University Hospital, Saitama, Japan

[10]Center for Medical Genetics, Chiba Children's Hospital, Chiba, Japan

[11]Laboratory for Comprehensive Genomic Analysis, RIKEN Center for Integrative Medical Sciences, Yokohama, Kanagawa, Japan

[12]These authors contributed equally

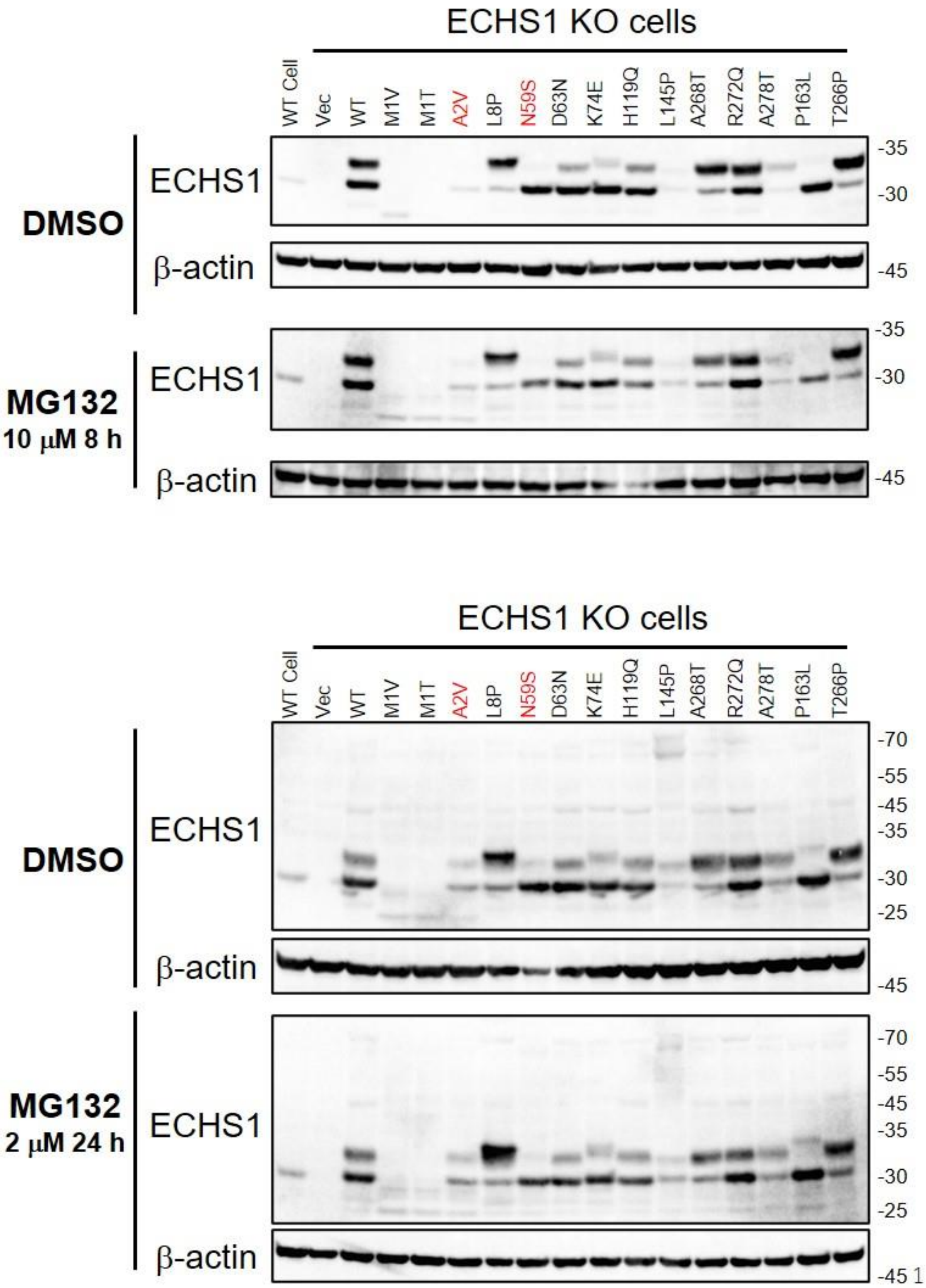


**Supplementary Figure 1. Protein stability of ECHS1 variants.**

WT and *ECHS1* KO HEK293FT cells were transfected with indicated expression vectors. 24 h after transfection, cells were treated with 10 μM MG132 for 8 h (A) or 2 μM MG132 for 24 h (B) and subjected to immunoblotting analysis using indicated antibodies.

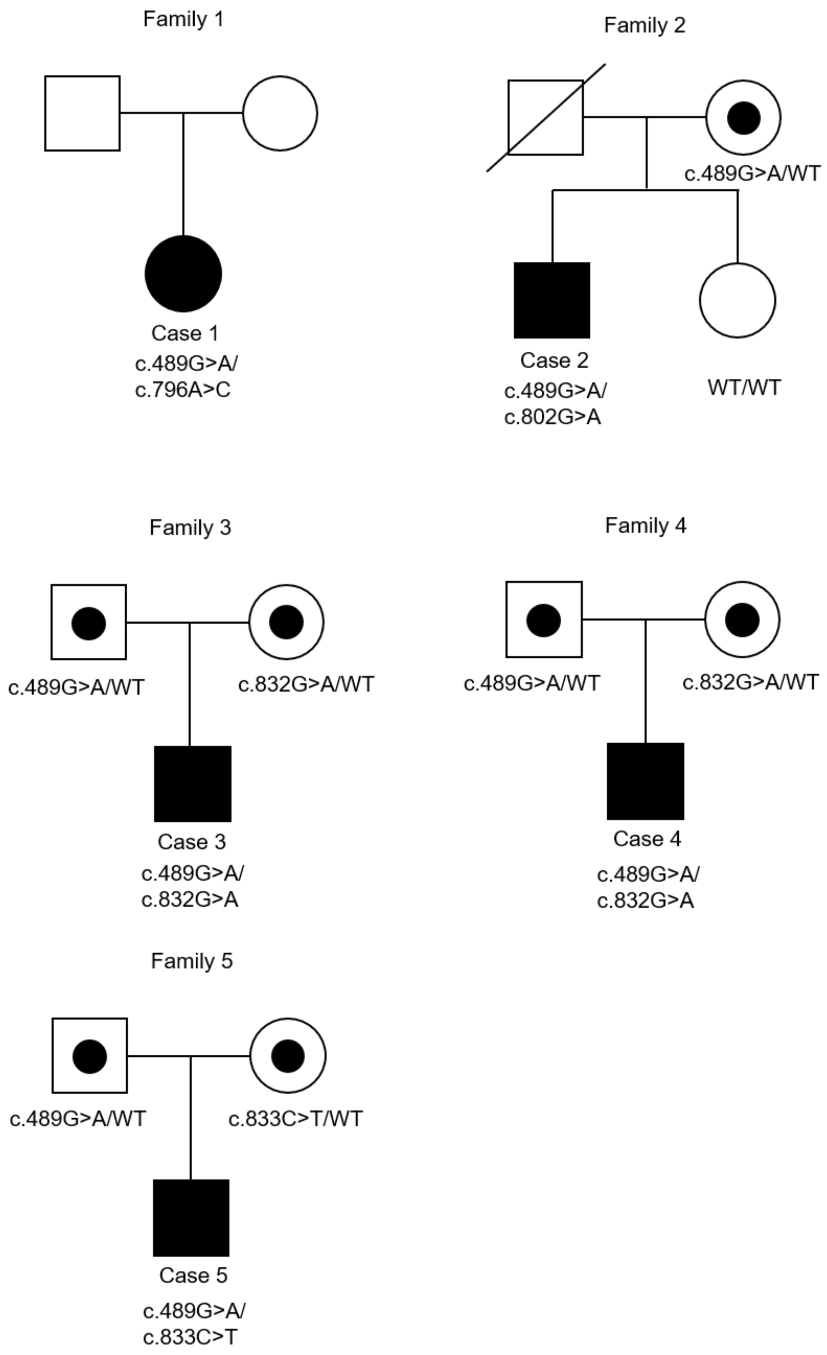


Supplemental Figure 2

**Supplementary Figure 2. Family pedigree for each family with ECHS1 variants**

Square and circles mean males and females, respectively. Dots represent carriers of the variant. Diagonal lines indicate deceased individuals. Symptomatic family members are shown in black filled. WT, wild type.

| NM_004092.4 | CDS_position | NP_004083.3 | Protein_position | Amino_acids | ClinVar |
|---|---|---|---|---|---|
| c.1A>G | 1 | p.Met1Val | 1 | M/V | Uncertain significance |
| c.2T>C | 2 | p.Met1Thr | 1 | M/T | - |
| c.5C>T | 5 | p.Ala2Val | 2 | A/V | Pathogenic |
| c.23T>C | 23 | p.Leu8Pro | 8 | L/P | Uncertain significance |
| c.98T>C | 98 | p.Phe33Ser | 33 | F/S | - |
| c.176A>G | 176 | p.Asn59Ser | 59 | N/S | Pathogenic |
| c.187G>A | 187 | p.Asp63Asn | 63 | D/N | Uncertain significance |
| c.220A>G | 220 | p.Lys74Glu | 74 | K/E | - |
| c.357C>G | 357 | p.His119Gln | 119 | H/Q | - |
| c.413C>T | 413 | p.Ala138Val | 138 | A/V | Pathogenic |
| c.434T>C | 434 | p.Leu145Pro | 145 | L/P | - |
| c.476A>G | 476 | p.Gln159Arg | 159 | Q/R | Pathogenic/Likely pathogenic |
| c.488C>T | 488 | p.Pro163Leu | 163 | P/L | Likely pathogenic |
| c.796A>C | 796 | p.Thr266Pro | 266 | T/P | - |
| c.802G>A | 802 | p.Ala268Thr | 268 | A/T | - |
| c.815G>A | 815 | p.Arg272Gln | 272 | R/Q | Uncertain significance |
| c.832G>A | 832 | p.Ala278Thr | 278 | A/T | Conflicting interpretations of pathogenicity |
| c.833C>T | 833 | p.Ala278Val | 278 | A/V | Uncertain significance |

**Supplementary Table 1. Detailed information about *ECHS1* variants**

In silico prediction information from dbNSFPv4.2, Pubmed article information, allele frequencies in gnomAD and jMorp, and variant interpretation in ClinVar are listed. D: damaging/deleterious, T: tolerated, B: benign, N: neutral, U: unknown, H: high, M: medium, L: low

| NM_004092.4 | gnomAD_exomes_AF | gnomAD_genomes_AF | jMorp(ToMMo 4.7KJPN) | jMorp(ToMMo 38KJPN) | CADD_phred |
|---|---|---|---|---|---|
| c.1A>G | 8.12E-05 | - | - | 0.000129 | 20.6 |
| c.2T>C | 1.02E-05 | 6.58E-06 | - | 0.000336 | 22.6 |
| c.5C>T | 1.02E-05 | 3.95E-05 | - | 0.00009 | 21.4 |
| c.23T>C | - | - | 0.0001 | 0.00009 | 22.7 |
| c.98T>C | - | - | - | - | 24.9 |
| c.176A>G | 3.6E-05 | 2.63E-05 | 0.0016 | 0.002311 | 24.2 |
| c.187G>A | 2.8E-05 | 2.63E-05 | 0.0003 | 0.000155 | 3.307 |
| c.220A>G | - | - | 0.0001 | 0.000039 | 11.14 |
| c.357C>G | 3.98E-05 | 2.63E-05 | 0.0003 | 0.000168 | 18.25 |
| c.413C>T | - | - | - | - | 32 |
| c.434T>C | - | - | 0.0001 | 0.000013 | 32 |
| c.476A>G | 0.000108 | 0.000112 | - | - | 22.8 |
| c.488C>T | 3.59E-05 | 1.97E-05 | - | - | 26.7 |
| c.796A>C | - | - | - | - | 31 |
| c.802G>A | - | - | - | - | 26.5 |
| c.815G>A | 3.98E-05 | 0.000125 | - | - | 19.4 |
| c.832G>A | 7.95E-06 | 1.97E-05 | - | - | 23.9 |
| c.833C>T | - | 6.57E-06 | - | - | 24.4 |

Supplementary Table 1 (continued)

| NM_004092.4 | Pred_Damaging/Deleterious (Of 16 in silico tools) | MetaLR_pred | MetaLR_score | MetaSVM_pred | MetaSVM_score | MetaRNN_pred | MetaRNN_score |
|---|---|---|---|---|---|---|---|
| c.1A>G | 6 | T | 0.1407 | T | -0.9301 | D | 0.872063 |
| c.2T>C | 7 | T | 0.1543 | T | -0.8961 | D | 0.8846 |
| c.5C>T | 3 | T | 0.1181 | T | -0.9637 | D | 0.714376 |
| c.23T>C | 4 | T | 0.1046 | T | -1.0346 | T | 0.457327 |
| c.98T>C | 6 | T | 0.0999 | T | -1.0864 | D | 0.818875 |
| c.176A>G | 13 | D | 0.8962 | D | 0.9525 | D | 0.930062 |
| c.187G>A | 1 | T | 0.1181 | T | -0.9878 | T | 0.049394 |
| c.220A>G | 0 | T | 0.0694 | T | -1.0157 | T | 0.069017 |
| c.357C>G | 3 | T | 0.1191 | T | -0.9726 | T | 0.208476 |
| c.413C>T | 10 | T | 0.4896 | D | 0.0848 | D | 0.984888 |
| c.434T>C | 13 | D | 0.7636 | D | 0.8533 | D | 0.960719 |
| c.476A>G | 4 | T | 0.1214 | T | -0.9679 | D | 0.510915 |
| c.488C>T | 13 | D | 0.6653 | D | 0.5462 | D | 0.970228 |
| c.796A>C | 10 | T | 0.4325 | T | -0.1833 | D | 0.811351 |
| c.802G>A | 7 | T | 0.3594 | T | -0.3156 | T | 0.45995 |
| c.815G>A | 4 | T | 0.1491 | T | -0.7925 | T | 0.100618 |
| c.832G>A | 10 | T | 0.4646 | D | 0.0907 | D | 0.92675 |
| c.833C>T | 8 | T | 0.4063 | T | -0.1148 | D | 0.940182 |

Supplementary Table 1 (continued)

| NM_004092.4 | BayesDel_addAF_pred | BayesDel_addAF_score | BayesDel_noAF_pred | BayesDel_noAF_score | DEOGEN2_pred | DEOGEN2_score |
|---|---|---|---|---|---|---|
| c.1A>G | T | 0.022513 | D | 0.112635 | T | 0.026987 |
| c.2T>C | D | 0.625005 | D | 0.66 | T | 0.018775 |
| c.5C>T | T | -0.0425 | T | -0.29883 | T | 0.041313 |
| c.23T>C | T | -0.0646 | T | -0.33057 | T | 0.031133 |
| c.98T>C | T | -0.14026 | T | -0.43924 | T | 0.180161 |
| c.176A>G | T | -0.0214 | D | -0.00409 | T | 0.307559 |
| c.187G>A | T | -0.44134 | T | -0.60731 | T | 0.071005 |
| c.220A>G | T | -0.24152 | T | -0.5847 | T | 0.032887 |
| c.357C>G | T | -0.25065 | T | -0.25509 | T | 0.133758 |
| c.413C>T | D | 0.35444 | D | 0.271352 | T | 0.145167 |
| c.434T>C | D | 0.427082 | D | 0.375697 | T | 0.331652 |
| c.476A>G | T | -0.11053 | T | -0.09622 | T | 0.052278 |
| c.488C>T | D | 0.273603 | D | 0.352878 | T | 0.183811 |
| c.796A>C | D | 0.321772 | D | 0.224427 | T | 0.141923 |
| c.802G>A | T | 0.021748 | T | -0.20654 | T | 0.121276 |
| c.815G>A | T | -0.34154 | T | -0.38564 | T | 0.121432 |
| c.832G>A | T | -0.02904 | T | -0.09797 | T | 0.135067 |
| c.833C>T | T | 0.024151 | T | -0.20309 | T | 0.135067 |

Supplementary Table 1 (continued)

| NM_004092.4 | FATHMM_pred | FATHMM_score | LIST-S2_pred | LIST-S2_score | LRT_pred | LRT_score |
|---|---|---|---|---|---|---|
| c.1A>G | T | 0.19 | D | 0.9 | - | - |
| c.2T>C | T | 0.1 | D | 0.9 | - | - |
| c.5C>T | T | -0.02 | T | 0.487551 | - | - |
| c.23T>C | T | -0.02 | T | 0.40306 | U | 0.374416 |
| c.98T>C | T | 1 | T | 0.551245 | N | 0.14286 |
| c.176A>G | D | -2.55 | D | 0.9957 | D | 0 |
| c.187G>A | T | -0.3 | D | 0.867413 | N | 0.033537 |
| c.220A>G | T | -0.14 | T | 0.841716 | N | 0.354445 |
| c.357C>G | T | -0.26 | T | 0.464953 | D | 0 |
| c.413C>T | T | 0.15 | D | 0.985868 | D | 0.000002 |
| c.434T>C | T | -0.83 | D | 0.918108 | D | 0.000004 |
| c.476A>G | T | -0.24 | D | 0.954805 | D | 0 |
| c.488C>T | T | -0.69 | D | 0.997267 | D | 0 |
| c.796A>C | T | -0.25 | D | 0.952805 | D | 0 |
| c.802G>A | T | -0.3 | D | 0.918808 | D | 0 |
| c.815G>A | T | -0.3 | D | 0.952405 | N | 0.00465 |
| c.832G>A | T | 0.43 | D | 0.987868 | D | 0.000005 |
| c.833C>T | T | 0.44 | D | 0.938806 | N | 0.000005 |

Supplementary Table 1 (continued)

| NM_004092.4 | M-CAP_pred | M-CAP_score | MutationAssessor_pred | MutationAssessor_score | MutationTaster_pred | MutationTaster_score |
|---|---|---|---|---|---|---|
| c.1A>G | D | 0.941035 | - | - | D | 1 |
| c.2T>C | D | 0.957347 | - | - | D | 1 |
| c.5C>T | D | 0.316741 | N | 0.55 | N | 0.998309 |
| c.23T>C | D | 0.217802 | L | 1.04 | N | 0.999999 |
| c.98T>C | T | 0.007368 | L | 1.04 | D | 0.964231 |
| c.176A>G | D | 0.303161 | H | 4.26 | D | 0.999997 |
| c.187G>A | T | 0.007115 | L | 1.095 | N | 0.986898 |
| c.220A>G | T | 0.006232 | N | -1.11 | N | 0.999971 |
| c.357C>G | T | 0.007999 | N | -0.185 | D | 0.999993 |
| c.413C>T | T | 0.018576 | M | 2.46 | D | 1 |
| c.434T>C | D | 0.241285 | H | 3.645 | D | 1 |
| c.476A>G | T | 0.006954 | N | -1.255 | D | 0.999997 |
| c.488C>T | D | 0.096996 | M | 3.21 | D | 1 |
| c.796A>C | D | 0.093958 | L | 1.71 | D | 1 |
| c.802G>A | D | 0.052732 | L | 1.2 | D | 0.999926 |
| c.815G>A | T | 0.020844 | N | 0.1 | D | 0.999931 |
| c.832G>A | D | 0.030457 | H | 3.61 | D | 0.999997 |
| c.833C>T | D | 0.061124 | H | 4.415 | D | 0.999995 |

Supplementary Table 1 (continued)

| NM_004092.4 | PROVEAN_pred | PROVEAN_score | PolyPhen_pred | PolyPhen_score | PrimateAI_pred | PrimateAI_score | SIFT_pred | SIFT_score |
|---|---|---|---|---|---|---|---|---|
| c.1A>G | N | -1.36 | B | 0.186 | - | - | D | 0.05 |
| c.2T>C | N | -1.22 | B | 0.419 | - | - | D | 0 |
| c.5C>T | N | -1.64 | B | 0.011 | D | 0.869824 | T | 0.08 |
| c.23T>C | N | -1.58 | D | 0.497 | D | 0.830297 | D | 0.01 |
| c.98T>C | D | -5.18 | D | 0.697 | D | 0.804998 | D | 0.01 |
| c.176A>G | D | -4.69 | D | 0.992 | T | 0.590744 | D | 0 |
| c.187G>A | N | -0.5 | B | 0.003 | T | 0.274067 | T | 0.51 |
| c.220A>G | N | 1.76 | B | 0.001 | T | 0.353441 | T | 0.69 |
| c.357C>G | D | -4.76 | B | 0.015 | T | 0.434698 | T | 0.2 |
| c.413C>T | D | -3.76 | D | 0.997 | T | 0.593288 | D | 0 |
| c.434T>C | D | -5.94 | D | 1 | T | 0.668584 | D | 0 |
| c.476A>G | N | -1.24 | B | 0.005 | T | 0.723277 | T | 0.23 |
| c.488C>T | D | -9.59 | D | 1 | D | 0.803635 | D | 0 |
| c.796A>C | D | -4.84 | D | 0.978 | T | 0.560642 | D | 0.03 |
| c.802G>A | D | -3.32 | D | 0.808 | T | 0.618345 | D | 0 |
| c.815G>A | D | -2.95 | D | 0.757 | T | 0.401216 | T | 1 |
| c.832G>A | D | -3.77 | D | 0.985 | T | 0.523828 | D | 0 |
| c.833C>T | D | -3.72 | D | 0.862 | T | 0.544106 | D | 0.02 |

Supplementary Table 1 (continued)

| DNA (NM_004092.4) | Protein (NP_004083.3) | jMorp(ToMMo 4.7KJPN) | jMorp(ToMMo 38KJPN) | ClinVar |
|---|---|---|---|---|
| c.23T>C | p.Leu8Pro | 0.0001 | 0.00009 | |
| c.187G>A | p.Asp63Asn | 0.0003 | 0.000155 | |
| c.220A>G | p.Lys74Glu | 0.0001 | 0.000039 | |
| c.357C>G | p.His119Gln | 0.0003 | 0.000168 | |
| c.434T>C | p.Leu145Pro | 0.0001 | 0.000013 | |
| c.5C>A | p.Ala2Val | N.D. | 0.00009 | Pathogenic |
| c.176A>G | p.Asn59Ser | 0.0016 | 0.002311 | Pathogenic |
| c.10dupC | p.Leu4fs | N.D. | 0.000013 | |
| c.86C>T | p.Ser29Leu | N.D. | 0.000013 | |
| c.137C>T | p.Thr46Ile | N.D. | 0.000026 | |
| c.160C>T | p.Arg54Cys | N.D. | 0.000013 | Likely_pathogenic |
| c.244G>A | p.Val82Met | N.D. | 0.000065 | Uncertain_significance |
| c.268G>A | p.Gly90Arg | N.D. | 0.000013 | Uncertain_significance |
| c.325C>T | p.Gln109* | N.D. | 0.000013 | |
| c.326A>G | p.Gln109Arg | N.D. | 0.000013 | |
| c.357C>G | p.His119Gln | N.D. | 0.000168 | |
| c.407G>A | p.Gly136Asp | N.D. | 0.000013 | |
| c.434T>C | p.Leu145Pro | N.D. | 0.000013 | |
| c.458A>G | p.Tyr153Cys | N.D. | 0.000013 | Uncertain_significance |
| c.518C>T | p.Ala173Val | N.D. | 0.000077 | Pathogenic/Likely_pathogenic |
| c.523G>A | p.Gly175Ser | N.D. | 0.000013 | Likely_pathogenic |

| | | | | |
|---|---|---|---|---|
| c.541delC | p.Arg181fs | N.D. | 0.000013 | |
| c.647_652delAGACAC | p.Glu216_Leu218delinsVal | N.D. | 0.000013 | |
| c.665C>G | p.Ala222Gly | N.D. | 0.000013 | |
| c.759_762delAGAA | p.Gly255fs | N.D. | 0.000013 | |
| c.848G>A | p.Arg283Lys | N.D. | 0.000013 | |
| c.867C>G | p.Asp289Glu | N.D. | 0.000013 | Uncertain_significance |

**Supplementary Table 2. The registration status of ECHS1 variants in jMorp**
